# Systematic Review and Meta-analysis of Immunomodulator and Biologic Therapies for Treatment of Chronic Pouchitis

**DOI:** 10.1101/2021.01.19.21250059

**Authors:** Emi Khoo, Andrew Lee, Teresa Neeman, Yoon-Kyo An, Jakob Begun

## Abstract

**Background:** Pouchitis is a common complication after restorative proctocolectomy with ileal pouch-anal anastomosis (IPAA). Although antibiotics are the primary therapy for acute pouchitis, a proportion of patients developed chronic antibiotic-dependent pouchitis (CADP) or antibiotic-refractory pouchitis (CARP). The efficacy of second line immunomodulator and biologic therapies for chronic pouchitis remain undefined. We performed a systematic review and metanalysis of published studies to assess their efficacy.

**Method:** The online EMBASE database was searched for full-text articles describing the treatment of chronic pouchitis meeting our criteria. Post-induction clinical and endoscopic response and remission rates were extracted and combined for meta-analyses. The rate of treatment discontinuation and safety profiles were also assessed.

**Results:** A total of 21 full-text articles were included in this meta-analysis representing 491 patients. The overall clinical response rate was 49% with clinical remission rate of 34%. The overall endoscopic response and remission rates were 53% and 36% respectively. The safety profile of individual agents was reassuring, but vedolizumab appears to have a more favorable safety profile.

**Conclusion:** This review and meta-analysis identified the effectiveness of vedolizumab and ustekinumab in achieving clinical and endoscopic response in chronic pouchitis, with a reassuring safety profile. There is limited data regarding use of immunomodulators and no conclusion can be drawn. Further studies are required to define the comparative effectiveness of available treatments of CADP or CARP.

## Introduction

Ileal pouch-anal anastomosis (IPAA) is a restorative surgical intervention following proctocolectomy for management of severe refractory ulcerative colitis (UC).^1^ An IPAA may also be formed after proctocolectomy for other indications such as familial adenomatous polyposis or colonic high-grade dysplasia secondary to inflammatory bowel disease (IBD).^2^ Pouchitis is a common complication of IPAA and affects up to 20% of patients within the first year post pouch formation, and up to 40% at 5 years.^3^ The Crohn’s and Colitis Foundation of America Partners cohort reported an overall incidence of pouchitis in up to 79% of patients after IPAA for UC.^4^ Interestingly pouchitis is more common in patients who had an IPAA performed for UC than for other indications, indicating that a similar pathogenesis may underly the development of pouchitis and UC. Symptoms of pouchitis can be debilitating, including increased stool frequency, rectal bleeding, abdominal cramping, urgency and faecal incontinence.^5^ A diagnosis of pouchitis should be confirmed with endoscopy and mucosal biopsy of the pouch, in order to exclude other conditions including cuffitis and functional pouch disorder.^6^ Common findings on endoscopy include edematous mucosa, granularity, friability, loss of vascular pattern, mucous exudates and ulceration of pouch.^7^

The first line therapy for acute pouchitis is antibiotics, most commonly metronidazole and ciprofloxacin, which is highly effective.^11^ Up to 87.5% of patients treated with antibiotics achieved clinical remission, while 88% achieved clinical response.^1^ Similarly, significant improvements of median PDAI score have been described with antibiotic therapy, which was strongly correlated with the patients’ satisfaction and quality of life.^12^ However, up to 15% of patients develop chronic antibiotic-dependent pouchitis (CADP) or chronic antibiotic-resistant pouchitis (CARP).^13-14^ A diagnosis of CADP or CARP is established if patients require continuous antibiotic therapy for more than 4 weeks within a 12 month period to relieve symptoms, or fail to achieve an adequate response despite a 4 week course of antibiotic use respectively.

Immunomodulator and biologic therapies are often prescribed for management of chronic pouchitis. These medications target pro-inflammatory pathways that are believed to drive inflammation in chronic pouchitis. Evidence supporting their use is limited to retrospective cohort studies and pilot studies. High level comparative evidence on immunomodulator and biologic therapies for chronic pouchitis is needed to support a treatment algorithm for CARP and CADP. To investigate the available evidence for therapies in chronic pouchitis we performed a systemic review and meta-analysis to determine the efficacy of immunomodulator and biologic therapies for treatment of chronic pouchitis in adult patients who have undergone an IPAA for UC.

## Methods

Our study was conducted in accordance with the Preferred Items for Systematic Reviews and Meta-analysis [PRISMA] guidelines. Our systematic review was registered with the International prospective register of systematic reviews (PROSPERO) on 7^th^ August 2020 (reference number: CRD42020196627).

### Study Selection

A structured search of the EMBASE database was performed on 1^st^ Dec 2019, to identify all studies that assessed the efficacy of immunomodulators and/or biologic therapies in chronic pouchitis. The Medical Subject Heading (MeSH) terms used were “pouchitis”, “ulcerative colitis”, “therapy”, “immunomodulating agent”, “methotrexate”, “azathioprine”, “mercaptopurine”, “tioguanine”, “thioguanine derivatives”, “tumor necrosis factor inhibitor”, “infliximab”, “adalimumab”, “golimumab”, “vedolizumab” and “ustekinumab”.

Articles meeting the search criteria were screened by two authors (EK, AL). Shortlisted publications were reviewed independently by two authors (YA, JB). Discrepancies at either stage of screening were resolved by discussion. For the purpose of this study, inclusion criteria were: adult patients who have undergone IPAA for indication of UC; articles that were published in complete form in peer-reviewed journals, either prospective or retrospective studies; articles that reported at least 3 patients treated for chronic pouchitis.

Articles were excluded if they were review articles, published in abstract form only, non-English language articles, studies reporting less than three cases of chronic pouchitis and those that reported duplicate results. Those studies which reported outcomes of non-immunomodulator and non-biologic interventions, were also excluded.

### Data Extraction and Outcome Measures

Baseline characteristics, efficacy and safety were extracted from each eligible study. The primary outcome was the clinical response rate to immunomodulator or biologic therapy. Secondary outcomes included the clinical remission rate, endoscopic response rate, mucosal healing rate, and rate of discontinuation of immunomodulator or biologic therapy when reported. Clinical response and endoscopic response were defined as reduction of clinical and endoscopic sub score of PDAI by two or more, respectively. The definitions of clinical remission and mucosal healing were clinical and endoscopic sub scores of the PDAI of less than three. Reasons for discontinuation were extracted and included serious adverse events, surgical intervention and switch of biologic therapy due to treatment failure. It was anticipated that there would be variability in time points for assessment and follow up duration in each study. This review aimed to assess measured outcomes, at six months (with a four month window) following commencement of intervention. The reasons for discontinuation were reported at the end of follow up period.

### Grading of Evidence

Assessment of quality and risk of bias was completed by two independent authors (EK, AL), based on the Oxford Centre for Evidence-Based Medicine 2011 Levels of Evidence.^15^ The strength of evidence for each of the articles were judged based on the study design, assigning a level from 1 (high quality, low risk of bias) to 5 (low quality, high risk of bias). Any disagreement between two reviewers were resolved by discussion.

### Statistical Analysis

The overall response rates and 95% confidence intervals (CIs), including clinical response, clinical remission, endoscopic response and mucosal healing, were estimated using random intercept logistic regression models. This was performed using the *metaprop* function in the *meta* package (version 4.12-0) in R (version 3.6.3). For the rate of discontinuation, the events including IBD-related surgery, switch of therapy and serious adverse events were combined. Statistical heterogeneity described the percentage of total variation across studies that are attributable to between-study heterogeneity. This was determined using the Q statistic of I^2^. An I^2^ value of more than 50% represented significant statistical heterogeneity.

## Results

### Study selection

The EMBASE literature search identified a total of 1260 publications. After the initial screening based on paper abstracts, 44 full text articles were reviewed. Twenty-one articles were considered eligible for meta-analysis following the application of inclusion and exclusion criteria. (*Figure 1*).

**Figure 1.**
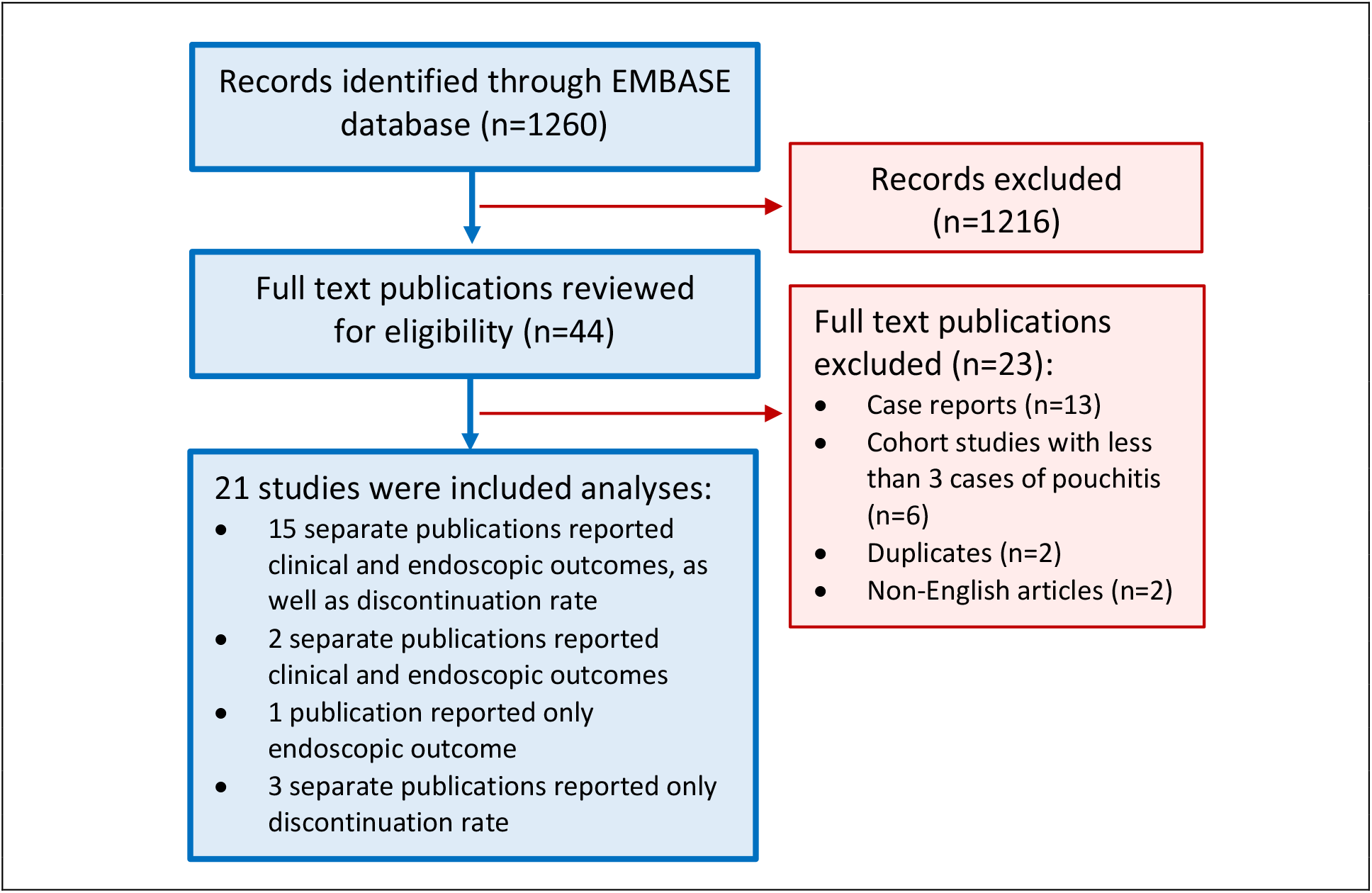
Inclusion of the studies for meta analyses of immunomodulator and biologic therapies in chronic pouchitis.

### Grading of Evidence

Thirteen studies were graded as level 4 strength of evidence and eight studies were graded as level 3 (*Table 1*). This included a single randomized controlled trial for which the strength of evidence was graded as 3 on the basis of small absolute effect size.

**Table 1.** Study characteristics and patient demographics from studies included in systemic literature review

| Study | Types of study | Intervention | Total cases | Mean age | Female | Median follow up (month) | Clinical response | Clinical remission | Endoscopic response | Endoscopic remission | Discontinuation | Assessment of bias |
| --- | --- | --- | --- | --- | --- | --- | --- | --- | --- | --- | --- | --- |
| Ollech et al. 2019 | Retrospective | Ustekinumab | 24 | 35.6 | 10 (41.7%) | 13 | 50.0% | - | 38.5% | 0.0% | 20% | 4 |
| Weaver et al. 2019 | Retrospective | Ustekinumab | 56 | 44.1 | 21 (37.5%) | 12 | 62.5% | 7.1% | 60.0% | 0.0% | 13% | 4 |
| Gregory et al. 2019 | Retrospective | Vedolizumab | 83 | - | 45 (54.2%) | 15.5 | 41.0% | 16.9% | 55.4% | 14.9% | 36% | 4 |
| Singh et al. 2019 | Retrospective | Vedolizumab | 19 | 26.7 | 9 (47.4%) | 3 | 31.6% | 0.0% | 73.7% | - | 21% | 4 |
| Verstockt et al. 2019 | Retrospective | Vedolizumab | 15 | 49.7 | 5 (33.3%) | 14.5 | - | 60.0% | - | - | 27% | 4 |
| Bar et al. 2018 | Retrospective | Vedolizumab | 20 | 22.5 | 8 (40.0%) | 3.5 | - | 45.0% | - | 64.3% | 0% | 4 |
| Khan et al. 2018 | Retrospective | Vedolizumab | 12 | 41.0 | 9 (75.0%) | 6 | 66.7% | 0.0% | 83.3% | 0.0% | - | 4 |
| Philpott et al. 2017 | Retrospective | Vedolizumab | 4 | 53.8 | 3 (75.0%) | 4 | - | - | 75.0% | 0.0% | - | 4 |
| Verstockt et al. 2019 | Retrospective | Infliximab | 23 | 41.0 | 6 (26.1%) | 14.5 | - | 43.5% | - | - | 74% | 4 |
| Segal et al. 2018 | Retrospective | Infliximab | 34 | 25.0 | 15 (44.1%) | 10 | - | - | - | - | 53% | 4 |
| Kelly et al. 2016 | Retrospective | Infliximab | 42 | 32.6 | 28 (66.6%) | 24 | 73.8% | 47.6% | - | 45.2% | 38% | 4 |
| Uchino et al. 2015 | Pilot | Infliximab | 10 | 23.0 | 7 (70.0%) | 2 | - | - | - | - | 30% | 3 |
| Viazis et al. 2013 | Pilot | Infliximab | 7 | 37.1 | 4 (57.1%) | 12 | 14.3% | 71.4% | - | 71.4% | 14% | 3 |
| Barreiro-De Acosta et al. 2012 | Retrospective | Infliximab | 33 | 45.0 | 15 (45.5%) | 12 | 33.3% | 33.3% | - | 40.0% | 39% | 4 |
| Haverman et al. 2011 | Retrospective | Infliximab | 4 | - | - | 38 | - | - | - | - | 25% | 4 |
| Ferrante et al. 2010 | Pilot | Infliximab | 25 | - | 14<br>(56.0%) | 21 | 56.0% | 32.0% | 15.4% | 61.5% | 48% | 3 |
| Calabrese et al. 2008 | Pilot | Infliximab | 10 | 39.2 | 4<br>(40.0%) | 6 | - | 80.0% | - | 80.0% | - | 3 |
| Viscido et al. 2003 | Pilot | Infliximab | 7 | 30.0 | 4<br>(57.1%) | 35 | 14.3% | 85.7% | 28.6% | 71.4% | 0% | 3 |
| Kjaer et al. 2019 | Randomised control trial | Adalimumab | 6 | 40.1 | 6<br>(100.0%) | 3 | 100.0% | 16.7% | 66.7% | - | 17% | 2 |
| Shen et al. 2009 | Pilot | Adalimumab | 17 | 36.0 | 5<br>(29.4%) | 2 | 23.5% | 47.1% | 23.5% | 41.1% | 18% | 3 |
| Verstockett et al. 2019 | Retrospective | Adalimumab | 13 | 39.3 | 5<br>(38.4%) | 14.5 | - | 38.5% | - | - | 77% | 4 |
| Barreiro-De Acosta et al. 2012 | Retrospective | Adalimumab | 10 | 43.2 | 3<br>(30.0%) | 12 | 30.0% | 10.0% | 0.0% | 40.0% | 40% | 4 |
| Uchino et al. 2013 | Pilot | Tacrolimus | 10 | 39.0 | 2<br>(20.0%) | 2 | 90.0% | 70.0% | 100.0% | 0.0% | 0% | 4 |
| Haverman et al. 2011 | Retrospective | Thiopurine | 8 | - | - | 38 | - | - | - | - | 13% | 4 |
| Haverman et al. 2011 | Retrospective | Infliximab + Thiopurine | 9 | - | - | 38 | - | - | - | - | 56% | 4 |

### Study characteristics

A total of 21 studies were included in this analysis; 13 retrospective studies, 7 cohort studies, and 1 randomized controlled trial. Thirteen of the 21 studies defined chronic pouchitis at baseline as the prolonged use of antibiotics for 4 weeks or more; while the remainder defined it as ongoing symptoms despite antibiotic use. Endoscopic confirmation of pouchitis at baseline was performed in 16 studies. Interventions included in the meta-analysis were ustekinumab (2 articles), vedolizumab (6 articles), infliximab (10 articles), adalimumab (4 articles), tacrolimus enema (1 article), thiopurine (1 article) and combination therapy of infliximab and thiopurine (1 article). All therapies were administered at recommended standard dosing regimens for ulcerative colitis.

A total of 491 patients from these studies were pooled for analysis, where the largest two cohorts were treated with either infliximab (n=185), or vedolizumab (n=153) (*Table 1*). Approximately 50% (n=247) of patients were female, with age ranging from 22 to 54 years. The reported age of pouch at commencement of intervention ranged from 1 to 10 years, based upon available data reported in 13 studies. The mean follow-up period ranged from 2 to 12 months.

Fifteen articles assessed clinical and endoscopic outcome, as well as discontinuation rate, 2 articles reported only clinical and endoscopic outcomes, another article reported only the endoscopic outcome, and 3 articles reported solely on the discontinuation rate (*Figure 1*).

### Primary Outcome

#### Clinical Response

Clinical response was assessed in 14 studies (Table 1 and Figure 2a), with the infliximab cohort making up the largest proportion of the sample, followed by vedolizumab, adalimumab, ustekinumab and lastly, tacrolimus. The overall pooled clinical response rate was 49% (95% CI, 36-62%; 14 studies, 175/351 patients). There was statistically significant between-study heterogeneity; *I*^2^ value of 79% (*P* > 0.01). Clinical response was achieved in 41% of patients who received infliximab (95% CI 21-65%); 42% of vedolizumab (95% CI 33-51%), 55% of adalimumab (95% CI 9-93%), 59% of ustekinumab (95% CI 48-69%) and 90% of tacrolimus (95% CI 53-99%). Between-study heterogeneity was evident only for infliximab treatment clinical response analyses (*I*^2^ = 77%; *P* < 0.01). The other interventional cohort studies, including ustekinumab, vedolizumab and adalimumab, showed low to moderate between-study heterogeneity.

**Figure 2.**
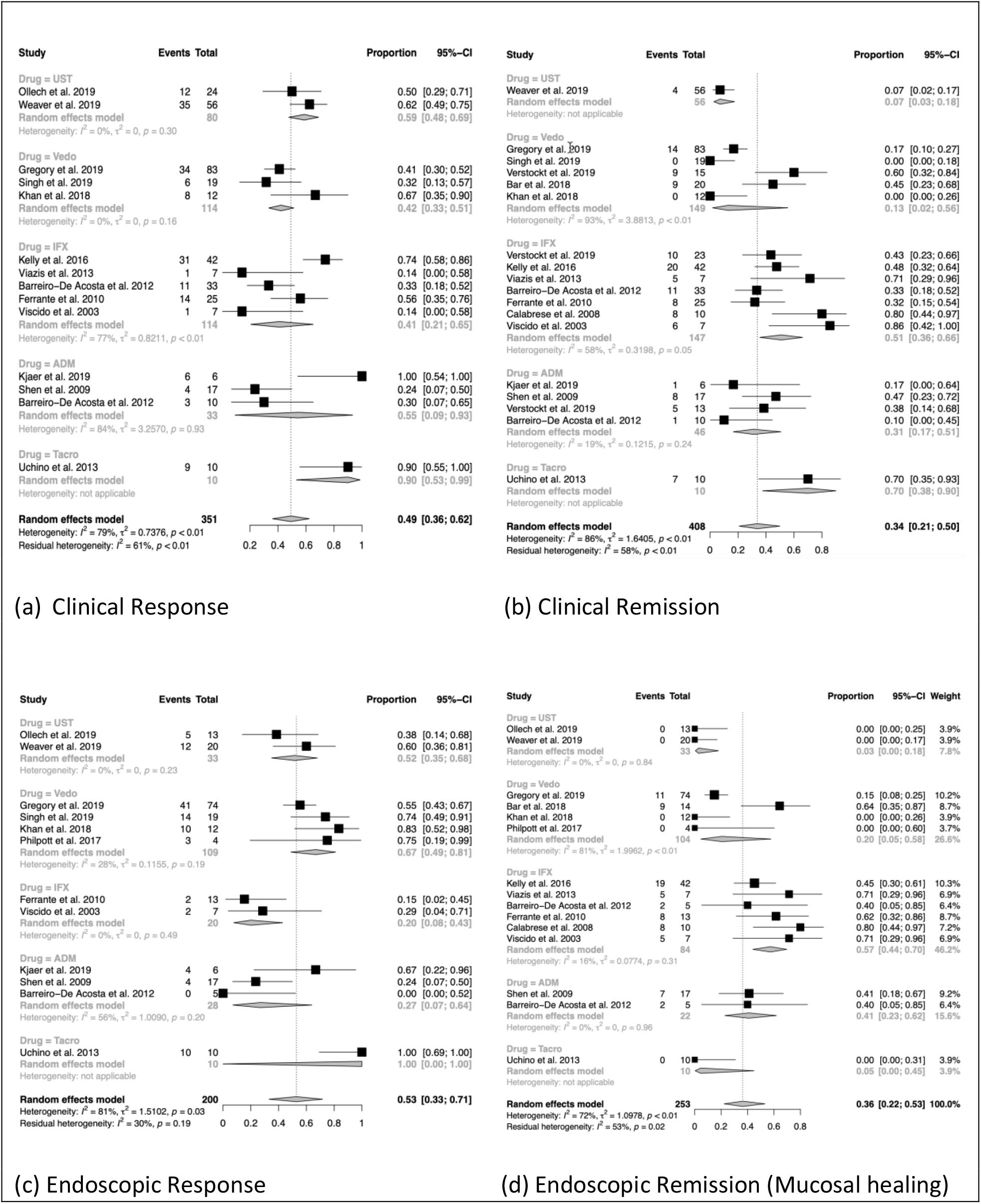
Primary and secondary outcomes comparing the efficacy of each interventions

### Secondary Outcomes

#### Clinical Remission

Eighteen studies reported on rates of clinical remission (Table 1 and Figure 2b). The majority of reported patients (n=408) received infliximab (n=149, 37%) or vedolizumab (n=147, 36%); while adalimumab, ustekinumab and tacrolimus made up a small proportion of the total cohort. The pooled clinical remission rate for immunomodulator and biologic therapies was 34% (95% CI 21-50%) with a high degree of between-study heterogeneity (*I*^2^ = 86%, *P* < 0.01). Clinical remission was observed in 51% of patient treated with infliximab (95% CI 36-66%), 13% of patient treated with vedolizumab (95% CI 2-56%), 31% treated with adalimumab (95% CI 17-51%), 7% treated with ustekinumab (95% CI 3-18%) and 70% of patients treated with tacrolimus (95% CI 38-90%). Low to moderate between-study heterogeneity was found in the infliximab and adalimumab cohorts, while between-study heterogeneity was found to be moderate to high in the vedolizumab cohort (*I*^2^ = 93%; *P* < 0.01).

#### Endoscopic Response

Twelve studies evaluated endoscopic response (Table 1 and Figure 2c). The overall endoscopic response rate for these studies was 53% (95% CI 33-71%). There was substantial between-study heterogeneity with *I*^2^ value of 81% (*P* = 0.03). Patients receiving vedolizumab accounted for more than half (n=109, 55%) of the pooled cohort analysed for endoscopic response rate with an individual endoscopic response rate of 67% (95% CI 49-81%). The remainder of the pooled cohort consisted of patients receiving ustekinumab, infliximab, adalimumab and tacrolimus in order of decreasing sample size. The endoscopic response rate for these groups was 52% (95% CI 35-68%), 20% (95% CI 8-43%), 27% (95% CI 7-64%) and 100% (95% CI 0-100%) respectively.

#### Endoscopic Remission

Fifteen studies reported endoscopic remission (Table 1 and Figure 2d). The combined results demonstrated a pooled endoscopic remission rate of 36% (95% CI 22-53%) with significant heterogeneity; *I*^2^ value of 72% (*P* < 0.01). Vedolizumab therapy contributed to the majority of the sample population with endoscopic remission rate reported. Endoscopic remission was was seen in 57% of patient treated with infliximab (95% CI 44-70%), 20% treated with vedolizumab (95% CI 5-58%), 41% treated with adalimumab (95% CI 23-62%), 3% treated with ustekinumab (95% CI 0-18%) and 5% treated with tacrolimus (95% CI 0-45%). There was low to moderate between-study heterogeneity reported in most studies, with an *I*^2^ of 0-16%, except for those vedolizumab studies endoscopic remission analyses (*I*^2^ = 81%; *P* < 0.01).

#### Discontinuation of therapy

Eighteen studies documented the rate of discontinuation at the end of follow up period, ranging from 2 to 38 months (Table 2). The highest rate of discontinuation was reported in cohorts receiving adalimumab (up to 77%), followed by infliximab (up to 74%), and the combination therapy of infliximab and thiopurine (56%). Ustekinumab and vedolizumab cohorts reported lower discontinuation rates of 13-20% and 0-36% respectively. One cohort study reported discontinuation rates of tacrolimus as 0% and thiopurine as 13%. The main reason for discontinuation was lack of response.

**Table 2.** Reasons of discontinuation of therapy in real world cohort of chronic pouchitis.

| Study | Intervention | Total cases | Median follow up (month) | Discontinuation (n, %) | Loss of response (n) | Switch of biologic therapy (n) | Surgical intervention (n) | Total adverse events (n, %) | Serious infection (n) | Serious adverse events (n) | Mild adverse events (n) |
| --- | --- | --- | --- | --- | --- | --- | --- | --- | --- | --- | --- |
| Ollech et al. 2019 | Ustekinumab | 24 | 13 | 5/24<br>20% | - | 3 | 2<br>8% | - | - | - | - |
| Weaver et al. 2019 | Ustekinumab | 56 | 12 | 7/56<br>13% | 6 | - | - | 1/56<br>2% | 1 | 0 | 0 |
| Gregory et al. 2019 | Vedolizumab | 83 | 15.5 | 30/83<br>36% | - | 0 | 30<br>36% | 5/83<br>6% | 3 | 0 | 2 |
| Singh et al. 2019 | Vedolizumab | 19 | 3 | 4/19<br>21% | - | 0 | 4<br>21% | 2/19<br>11% | 0 | 0 | 2 |
| Verstockett et al. 2019 | Vedolizumab | 15 | 14.5 | 4/15<br>27% | 4 | 1 | 0 | 0/15<br>0% | 0 | 0 | 0 |
| Bar et al. 2018 | Vedolizumab | 20 | 3.5 | 0/20<br>0% | 0 | 0 | 0 | 3/20<br>15% | 0 | 0 | 3 |
| Verstockett et al. 2019 | Infliximab | 23 | 14.5 | 17/23<br>74% | 8 | 13 | 4<br>17% | 9/23<br>39% | 0 | 1 | 8 |
| Segal et al. 2018 | Infliximab | 34 | 10 | 18/34<br>53% | 18 | 8 | 10<br>29% | 4/34<br>12% | 4 | 0 | 0 |
| Kelly et al. 2016 | Infliximab | 42 | 24 | 16/42<br>38% | 5 | 7 | 9<br>21% | 5/42<br>12% | 0 | 4 | 1 |
| Uchino et al. 2015 | Infliximab | 10 | 2 | 3/10<br>30% | 3 | 0 | 3<br>30% | - | - | - | - |
| Viazis et al. 2013 | Infliximab | 7 | 12 | 1/7<br>14% | 1 | 0 | 1<br>14% | 1/7<br>14% | 0 | 0 | 1 |
| Barreiro-de-Acosta et al. 2012 | Infliximab | 33 | 12 | 13/33<br>39% | 13 | 0 | 4<br>12% | 5/33<br>21% | 0 | 1 | 4 |
| Haverman et al. 2011 | Infliximab | 4 | 38 | 1/4<br>25% | 1 | 0 | 1<br>25% | 0/4<br>0% | 0 | 0 | 0 |
| Ferrante et al. 2010 | Infliximab | 25 | 21 | 12/25<br>48% | 9 | 0 | 6<br>24% | 2/25<br>8% | 0 | 2 | 0 |
| Viscido et al. 2003 | Infliximab | 7 | 35 | 0/7<br>0% | 0 | 0 | 0 | 1/7<br>14% | 1 | 0 | 0 |
| Kjaer et al. 2019 | Adalimumab | 6 | 3 | 1/6<br>17% | - | - | - | 0/6<br>0% | 0 | 0 | 0 |
| Shen et al. 2009 | Adalimumab | 17 | 2 | 3/17<br>18% | - | 0 | 3<br>18% | 4/17<br>24% | 0 | 0 | 4 |
| Verstocket et al. 2019 | Adalimumab | 13 | 14.5 | 10/13<br>77% | 6 | 4 | 2<br>15% | 2/13<br>15% | 0 | 2 | 0 |
| Barreiro-De Acosta et al. 2012 | Adalimumab | 10 | 12 | 4/10<br>40% | - | 0 | 4<br>40% | - | - | - | - |
| Uchino et al. 2013 | Tacrolimus | 10 | 2 | 0/10<br>0% | 0 | 0 | - | 3/10<br>30% | 0 | 0 | 3 |
| Haverman et al. 2011 | Thiopurine | 8 | 38 | 1/8<br>13% | 0 | 1 | 0 | 1/8<br>13% | 0 | 1 | 0 |
| Haverman et al. 2011 | Infliximab + Thiopurine | 9 | 38 | 5/9<br>56% | - | 0 | 5<br>56% | 0/9<br>0% | 0 | 0 | 0 |

The need for surgical interventions, including end ileostomy and anastomotic stricture dilatation, were observed. These procedures were more frequently seen in the cohort of patients treated with combination infliximab and thiopurine therapy (56%). Other biologic monotherapies, namely adalimumab, infliximab and vedolizumab, were found to have similar surgical interventional rates, in the range of 15-40%, 0-30% and 0-46% respectively. Surgical intervention was observed in 20% of the single ustekinumab study.

Adverse events were reported by most of the studies (15/20 articles). Serious infections, including *Clostridium difficile* infection, Norovirus, sepsis, intra-abdominal abscess and Shingles, were reported in 2% of ustekinumab cohort, 3.6% of vedolizumab cohort and 12% of infliximab cohort. Serious adverse events, in particular anaphylaxis and delayed hypersensitivity reaction, were only observed with infliximab (up to 10%) and adalimumab (15%). Pancreatitis was documented in 13% of patients in the thiopurine cohort. Mild adverse events, for example, headache, injection site reaction, nausea and rash, were most common in the infliximab intervention group, with the highest reported rate of 39%. Adalimumab was also found to have similar mild adverse events with a rate of up to 24%. Thirty percent of tacrolimus enema cohort reported mild burning in the pouch upon application. Low rates of mild adverse events were noted in the vedolizumab cohort (up to 15%) and none in the ustekinumab cohort.

## Discussion

Although antibiotics such as ciprofloxacin and metronidazole are effective in induction of remission for acute pouchitis^1^, a proportion develop CADP or CARP. Second line therapies including steroids, bismuth, elemental diet and faecal microbiota transplant failed to achieve significant rates of clinical remission in a recent meta-analysis.^1^ Effective treatments for chronic pouchitis are required given its significant burden not only on physical health and quality of life, but also due to the significant associated economic burden.^16^ Immunomodulator and biologic therapies for chronic pouchitis have been evaluated in several studies recently and this study reviews this data. To date, and to the best of the authors knowledge, this is the most comprehensive meta-analysis of real-world data from peer-reviewed full-text manuscripts, focusing on immunomodulator and biologic therapies in chronic pouchitis.

Many medical therapies have been investigated for the treatment of chronic pouchitis in the last decade. A systemic review by Segal et al. in 2017 examined 21 studies that assessed the efficacy of steroids, bismuth, elemental diet, tacrolimus and faecal microbiota transplantation.^1^ It concluded that none of these therapies achieved significant rates of response in chronic pouchitis.^1^ A Cochrane Review from 2019 examining 5 articles evaluating the efficacy of probiotics, bismuth enema, glutamine suppository, butyrate suppository and adalimumab for treatment of chronic pouchitis showed similar results.^6^

The pathogenesis of pouchitis remains uncertain. Acute pouchitis is believed to be driven by a bacterial etiology, explaining the responsiveness to antibiotic therapy.^17^ Many studies have suggested that the development of chronic pouchitis is multi-factorial, similar to inflammatory bowel disease itself. The proposed etiologies for development of chronic pouchitis include recurrence of ulcerative colitis, bacterial dysbiosis within the ileo-anal pouch, immune dysregulation, mucosal ischemia and oxygen-free radical injury, reduction in short-chain fatty acids and genetic predisposition.^18^ Histologically, ileal pouch mucosa shows colonic metaplasia with increased crypt cell proliferation and chronic inflammatory cell infiltrate within the lamina propria.^18^ Given that the histological features are analogous to ulcerative colitis, it is plausible that immunomodulators and biologic therapies used in induction and maintenance of ulcerative colitis would be beneficial in the setting of chronic pouchitis.

A single cohort study of 10 patients receiving tacrolimus enemas was included in this meta-analysis. The clinical response (90%), clinical remission (70%) and endoscopic response (100%) rates were found to be the highest among all immunomodulator and biologic interventions. The results of this study must be interpreted with caution due to the small sample size. Endoscopic remission or mucosal healing was only found in 5% of patients, suggesting that topical tacrolimus may be useful in inducing short term clinical response, but not necessarily mucosal healing. Although none of the patients discontinued therapy at the end of follow up period, 30% reported mild burning sensation upon local application and no patient was able to tolerate the applied tacrolimus for 10 minutes. No serious adverse events were documented supporting the relative safety of local tacrolimus application.

Thiopurine use in chronic pouchitis was reported in a retrospective study of 8 patients, which focused on the safety profile of therapy. Efficacy was not able to be assessed as clinical and endoscopic responses were not reported. After 38 months of follow-up, one male patient (13%) discontinued therapy due to acute pancreatitis. Thiopurine-induced pancreatitis is a well recognised complication occurring in approximately 3% of patients treated with thiopurines and is more common in females.^19^ The higher frequency noted in this study likely relates to the small sample size.

In our meta-analysis the largest cohort of patients was treated with infliximab and was associated with high clinical and endoscopic remission rates of 51% (95% CI 36-66%; *I*^2^ = 58%; 7 studies; 58/147 patients) and 57% (95% CI 44-70%; *I*^2^ = 16%; 6 studies; 47/84 patients) respectively, similar to previous results.^1^ Clinical response was reported in a proportion of studies and was achieved in 41% (95% CI 21-65%; *I*^2^ = 77%; 5 studies; 59/114 patients) of patients, however there was significant heterogeneity between studies. The reported endoscopic response rate was low at 20% (95% CI 8-43%; *I*^2^ = 0%; 2 studies; 4/20 patients). The contrast in rates of clinical and endoscopic response versus remission may reflect differences in available data as there was significant variability in outcomes reported amongst studies. The discontinuation rate in the infliximab cohort was noted to be as high as 74% in one of the retrospective studies. Reasons for discontinuation include non-response, loss of response and adverse events. Up to 30% of patients proceeded to formation of a permanent end-ileostomy at the end of follow up period. Almost 10% of patients suffered serious adverse events, including anaphylaxis, delayed hypersensitivity and drug-induced lupus. In addition, up to 12% of patients had severe infection, in particular, sepsis and Shingles. Overall infliximab was effective for clinical and endoscopic outcomes; however, the safety profile must be considered in patients receiving infliximab for CARP or CADP.

Similarly, adalimumab use in chronic pouchitis was associated with high rates of discontinuation and progression to surgical management, up to 77% and 40% respectively. A proportion of patients suffered adverse events, including delayed hypersensitivity (15%), headache and injection site reaction (24%). No serious infections were reported. This suggested a more tolerable safety profile for adalimumab, similar to previous published data.^20^ Overall clinical and endoscopic remission rate of adalimumab were noted to be 31% and 41% respectively, with low between-study heterogeneity. The lower rates of response with adalimumab may be due to differences in the patient populations reported, particularly the inclusion of patients who had previously failed infliximab in studies examining adalimumab. It is important to note that the one RCT included in this analysis was of adalimumab, and there was no significant difference in response between placebo and adalimumab arms, although this was a small study. In our analysis the adalimumab cohort had a reasonably high clinical response rate of 55%, but the large confidence interval (95% CI 9-93%) suggests that the true effect remains unclear. Taken together these results raise questions as to the efficacy of adalimumab for treatment of chronic pouchitis.

There were 6 full text articles assessing efficacy of vedolizumab in chronic pouchitis retrospectively, including patients who previously failed anti-tumor necrosis factor (anti-TNF) therapy. Clinical and endoscopic response rates were 42% and 67% respectively, without significant between-study heterogeneity. However clinical and endoscopic remission rates were poor at 13% and 20% respectively, with substantial between-study heterogeneity. This may reflect the slower onset of action of vedolizumab as well as a lack of correlation between endoscopic and clinical outcomes in pouchitis. The rate of discontinuation and surgical outcomes were both up to 36%, which is lower than seen with infliximab or adalimumab. With respect to the safety profile of vedolizumab, there were 3 cases of serious infection (6%), which included *C. difficile* infection, norovirus and intraabdominal sepsis. No serious adverse events were reported. Vedolizumab appears to achieve clinical and endoscopic response with a favourable safety profile. A similar safety profile to placebo was reported in post-marketing data with vedolizumab.^21^

Ustekinumab was recently approved for use in IBD, and there were only two published retrospective studies assessing the efficacy of ustekinumab on chronic pouchitis. A large proportion of patients (70%) were anti-TNF-experienced prior ustekinumab treatment. All participants received ustekinumab throughout the follow up period of the study (median follow up period of 12-13 months). The clinical response and endoscopic response rates were high at 59% and 52% respectively, with negligible between-study heterogeneity. In comparison to the published data on the efficacy of ustekinumab in ulcerative colitis at the end of induction phase, the clinical response rate in chronic pouchitis was similar but a lower endoscopic improvement rate was observed.^22-23^ However, clinical remission and endoscopic healing rates were quite low at 7% and 3% respectively. The rate of discontinuation was up to 20% due to loss of response and 8% progressed to a permanent end-ileostomy. There was a single case of *C. difficile* infection reported and no other serious adverse events. The percentage of serious infections was similar to the published data. These data suggest that ustekinumab is safely able to achieve clinical and endoscopic responses, but not necessarily remission during the follow-up period in the published studies.

This study provides an up to date comprehensive review and analysis of the available studies examining immunomodulator and biologic therapy for the treatment of chronic pouchitis. There are, however, several limitations to this study. Most studies included in this meta-analysis were single centred cohort studies with small sample sizes. Only one randomised placebo-controlled trial was included, however the small study population (n=6) compromises the quality of those results. The lack of high-quality head-to-head trials and the heterogeneity of the patient populations studied, makes it challenging to compare the benefit of one drug to another; and conclusions about the comparative efficacy of each agent cannot be drawn from the available data. For these reasons, data should be interpreted with caution. Through our analysis, we also found that inconsistent definitions of chronic pouchitis are used in the literature. The considerable between-study heterogeneity suggested the different disease activity measures used to assess response and remission, which impact the extrapolation of these findings to clinical practice. To overcome these limitations, large-scale multi-centre randomised placebo-controlled trials with a consensus definition of chronic pouchitis and the standardized outcome measures are required.

## Conclusion

Treatment of chronic antibiotic-refractory and antibiotic-dependent pouchitis remains challenging due to a lack of evidence guiding therapeutic decisions. This review and meta-analysis of real-world studies identified the effectiveness of vedolizumab and ustekinumab in achieving clinical and endoscopic response in chronic pouchitis, with a reassuring safety profile. Infliximab was found to be effective in inducing clinical and endoscopic remission, but there were significant rates of treatment failure and adverse events. Adalimumab demonstrated high rates of treatment discontinuation and progression to surgical outcomes despite reasonable clinical and endoscopic remission rates. There is limited published data regarding the use of immunomodulators for chronic pouchitis and no conclusion can be drawn. Further studies are required but current evidence favors vedolizumab, ustekinumab and infliximab for treatment of CADP or CARP.

## Data Availability

I confirmed that all data referred to in the manuscript is available.

